# Centenarians and Oldest Olds in Liguria – COOL: a multidisciplinary study to investigate the genetic determinants of cognitive well-being in Genoa, Italy. Rationale, study protocol and cohort profile

**DOI:** 10.64898/2026.05.20.26353506

**Authors:** Emilio Di Maria, Carlotta Gualco, Erika Muscolino, Nicoletta Reale, Claudio Marcello Solaro, Laura Camia, Umberto Tortorolo, Claudio Ivaldi, Luca Mazzella, Fabio Bandini, Elia Maioli, Manuela Stella, Francesca Mattioli, Erika Zumerle, Gaddo Flego, Michela Mazzocco, Nicoletta Sacchi, Angelo Schenone, Mauro Tettamanti, Gabriella Marcon, The COOL study Investigators, Massimo Del Sette

**Affiliations:** Department of Health Sciences, University of Genova, Genova, Italy; University Unit of Medical Genetics, Galliera Hospital, Genoa, Italy; Neurology Unit, Galliera Hospital, Genoa, Italy; Istituto Don Orione – Paverano, Genoa, Italy; Department of Geriatrics, Center for Cognitive Disorders and Dementia, ATS ASL 3, Genoa, Italy; Department of Neurology, Center for Cognitive Disorders and Dementia, ATS ASL 3, Italy; Unit of Internal Medicine, Villa Scassi Hospital, IRCCS AOM, Genoa, Italy; UOC Paediatric Clinical Trial Center, IRCCS Istituto Giannina Gaslini, Genoa, Italy; Department of Internal Medicine, Pharmacology & Toxicology Unit, University of Genoa, Genoa, Italy; Unit of Clinical Pharmacology, Galliera Hospital, Genoa, Italy; Ospedale Evangelico Internazionale, Genoa, Italy; Laboratorio di Istocompatibilità and Italian Bone Marrow Donor Registry – IBMDR, Galliera Hospital, Genoa, Italy; Department of Neurosciences, Rehabilitation, Ophthalmology, Genetic and Maternal Infantile Sciences, University of Genoa, Genoa, Italy; Neurology Clinic, IRCCS Azienda Ospedaliera Metropolitana Genovese, Genoa, Italy; Department of Health Policy, Istituto di Ricerche Farmacologiche Mario Negri IRCCS, Milan, Italy; Department of Medicine, University of Udine, Udine, Italy; Department of Medicine, Surgery and Health Sciences, University of Trieste; The COOL study Investigators are listed in the Collaborators section; Neurology Unit, IRCCS Azienda Ospedaliera Metropolitana Genovese, Genoa, Italy

**Keywords:** **MESH**, dementia, aged, cognition, neurogenetics, genetics, **KWs**, ageing, centenarians, protocol

## Abstract

**Objectives:** Despite the body of literature on genetic risk factors for dementia, little is known on protective genetic factors associated with favourable cognitive ageing in the oldest population. In Europe, Italy has a leading position with a swelling population of centenarians, and the urban area of Genoa in the Liguria region has one of the highest prevalence of centenarians. The COOL study is a not-for-profit, multicentric study involving a cohort of centenarians (aged >99) living in the Genoa area. The ultimate aim is the identification of genomic biomarkers associated with cognition in the oldest old population.

**Results:** Participants underwent a semi-structured interview on personal, disease and family history, and a neuropsychological assessment of the main cognitive domains. As of July 2025, we enrolled 88 centenarians (age range: 99-108, median 100.56) with and without cognitive impairment; 32 subjects were followed up. All participants were of Italian ancestry, 81% were female. The cognitive profile in assessed subjects showed a wide range of cognitive health measures (CDR 0-5; MMSE 3-30, median 24). Whole peripheral blood and DNA samples from 67 participants were stored.

**Conclusions:** We demonstrated that the protocol is feasible, and acceptable by participants and their families. A comprehensive phenotype dataset was established, and DNA samples were stored. Centenarians exhibited a broad spectrum of cognitive profiles, from preserved cognition to severe dementia. These findings will eventually allow to interpret the profiles of genomic variants as associated with variability of cognitive performance in centenarians. The molecular underpinnings of healthy cognitive ageing could inform health policy strategies in the general population.

**Highlights:**

- The study protocol was feasible and well-accepted by centenarians and their families.
- A comprehensive phenotype dataset was established, and DNA samples were collected and stored.
- Centenarians exhibited a broad spectrum of cognitive profiles, from preserved cognition to severe dementia.
- Genomic profiling will eventually enable the investigation of genetic variability underlying cognitive health.
- Missing data were present, consistent with the study design and the characteristics of the extremely aged cohort.

## Introduction

Extreme longevity has ever been a prominent subject for scientific curiosity. Only in last decades, yet, an oldest age has become a reliable perspective for a small but substantial fraction of populations. According to the United Nations (UN) Department of Economic and Social Affairs, Population Division, the global number of centenarians, as estimated from 1990 to 2024, increased by approximately two times every ten years (United Nations, 2024) – i.e., the global number of centenarians increases more and more rapidly over time (Kinsella and Phillips, 2005; Robine and Cubaynes, 2017). Though the number of centenarians is estimated to increase in diverse populations worldwide, large disparities exist among geographical areas. As a general view, the most affluent areas show the highest prevalence of oldest old (Robine and Cubaynes, 2017). This association may also explain the difference between urban areas and underserved, rural areas. Such disparities can be observed at all levels, even at national and regional scale. In Europe, the average is 17.3 centenarians per 100,000 inhabitants, with a striking difference (almost an order of magnitude) between the top countries and the bottom countries, the prevalence of centenarians ranging from ≤4 per 100,000 in Bulgaria and Romania, to 23 per 100,000 in Greece, 25.4 in Italy and 28.2 in France (Teixeira et al., 2017).

According to the latest analysis on centenarians released by the Italian National Institute of Statistics, as of January 1, 2025, there were 23,548 residents in Italy who were at least 100 years old, over 2,000 more than the previous year. Furthermore, this number has more than doubled (>130%) in the last fifteen years. Those aged 105 and over (named semi-supercentenarians) were 724, with the share of women rising to 90.7%; the supercentenarians aged >110 were 19, of whom only one is male (ISTAT, 2025).

In line with the general model for social determinants of health and disease, the complex interplay between constitutional factors and environmental factors occurs along the entire life span of the individuals. The genetic background is the fundamental non-modifiable, constitutional factor which influences health status in humans from conception to the end of life. Human longevity is in part genetically determined. Heritability of lifespan in adults was estimated to be greater than 0.2 in several cohorts of European descent (Herskind et al., 1996; Mitchell et al., 2001). Interestingly, heritability of longevity differs between man (0.26) and women (0.23) (Cournil et al., 2000), and increases with age (Murabito et al., 2012; Sebastiani and Perls, 2012).

Cognitive impairment is not an inherent feature of ageing, as it is acknowledged that it is possible to reach extreme ages with no sign of dementia (Qiu and Fratiglioni, 2018). Nonetheless, the increasing number of extremely old individuals is currently considered an emerging issue in public health, as global data suggest that the proportion of life in good health has remained broadly constant, implying that the additional years are in poor health (World Health Organization, 2020). This evolving public health scenario led to move the focus from longevity to healthy ageing (Khachaturian and Khachaturian, 2005; Menassa et al., 2023). The UN designated the period between 2021 and 2030 as the Decade of Healthy Aging to foster the development of programmes, strategies, and research aimed at reducing health inequities and improving the lives of older people, their families and communities (Eozenou et al., 2021; World Health Organization, 2020). Notably, as dementia is a major cause of morbidity and disability in the aged population, the age-related cognitive decline is a major issue for health and social services.

### Study rationale

The genetic background which counteracts the age-related cognitive decline is largely unexplored, as well as its interaction with life experience and environmental context. A favourable cognitive status in the elderly clearly correlates with longevity, but is a different phenotype – and, as such, the underlying determinants should be specifically addressed.

To date, most scientific efforts rather addressed the fundamental causes of neurodegeneration for which Alzheimer’s disease (AD) is the most frequent and clinically relevant model. The discovery of rare pathogenic variants in amyloid precursor protein (*APP*), presenilin 1 (*PSEN1*), and presenilin 2 (*PSEN2*) genes as causative of early-onset familial AD (which accounts for ∼1% of AD cases) provided important insights in the molecular mechanisms involved in AD pathogenesis (recently reviewed by Karagas et al., 2025). Then, starting from the seminal discovery of the *APOE* ε4 as a major risk allele (Corder et al., 1993), plenty of studies on large cohorts of individuals with sporadic AD allowed to discover additional, common genetic risk factors with low effect size (Karagas et al., 2025). However, even the relatively large effect on cognition determined by *APOE* ε4 is influenced by social support (Payen et al., 2024), thus confirming that environmental variables interact with genetic variability also in older adults.

Compared to the body of evidence on risk factors associated with increased susceptibility to AD, less is known about genetic factors underlying resistance and resilience to the disease (whereas resistance is the avoidance of AD neuropathologic changes, resilience is the ability to maintain cognitive function despite significant pathology (Arenaza-Urquijo and Vemuri, 2018) and, as such, has been recently highlighted as the key concept in dementia prevention strategies (Livingston et al., 2024). A handful of genetic protective variants were consistently identified so far. The relevant genes are mostly involved to the amyloid cascade (*APOE*, *CLU*, *PICALM*), tau pathology (*PLCG2*, *RELN*) or glial function (*ABCA7*, *TREM2*), according to a recent review (Marino et al., 2025).

The demonstration of AD pathology in extremely aged individuals without cognitive impairment (Zhang et al., 2023) invokes the mechanism of resilience in the oldest old population, underlining that risk factors for cognitive impairment in the oldest-old differ from younger elderly. The recent study from the Colombian centenarians reported that the major risk factors identified in adult midlife, such as smoking, body mass index and diabetes, do not influence the cognitive profile (Lozada-Martinez et al., 2025). The analyses on a large sample of individuals without dementia from the Netherlands demonstrated that vascular disorders do not influence the risk for dementia after ≥90 years of age (Legdeur et al., 2019). Our systematic review further confirmed that also the educational level, which is a well-established risk factor for dementia in the general population, does not correlate with the cognitive performance in the oldest-old individuals (Gualco et al., 2025).

It is also worth to note that the pathological hallmarks of neurodegeneration show different pattern in oldest individuals, as compared to the aged counterpart. A relevant proportion of dementia in the oldest-old is related to non-AD pathologies (Beker et al., 2021), while other causes of dementia are increasingly prevalent. Namely, the TDP-associated pathology reaches a peak close to 100 years of age (Kawas et al., 2021), whereas individuals who develop Alzheimer disease normally die at a younger age (Perls, 2004).

The lines of evidence briefly described herein point to a research question which has not been deeply explored so far, that is whether human genetics may help to explain the observed variability in the cognitive profile in the oldest age. Beyond the current knowledge on the role for genetic factors influencing cognitive decline in the elderly, further investigation is needed to identify novel genomic biomarkers as predictors of healthy cognitive ageing.

Selected cohorts of extremely aged individuals, namely aged 100 years or more, are invaluable models for exposure to a favourable combination of protective factors, both environmental and constitutional, that in turn are largely genetically determined. Genetic factors are not modifiable. However, including known genetic variants in prediction models may improve the accuracy in the identification of population fractions that deserve specific preventative and therapeutic strategies. Conversely, the reliable identification of individuals who are relatively resilient to cognitive decline may allow to avoid an excess of medicalisation in elder subjects, and the allocation of the relevant resources in at-risk individuals (Andersen, 2020).

To address this scientific subject, we embarked in a project aimed at investigating a cohort of centenarians who have been living in the same urban area and shared the same social environment during ageing. The ground assumptions were that a sample of extremely aged individuals can show a wide variability in the cognitive profile – that is the phenotype of interest – and that part of this variability may be explained by the genetic diversity within the sample cohort. To pilot the investigation of this hypothesis, we developed the study “Centenarians and oldest-olds in Liguria – COOL: Investigation on cognitive status in centenarians from Genova” – which is named COOL from hereafter.

We took advantage by being located in the Liguria region, which in the Italian map of longevity showed a high prevalence of centenarians, ranking in the second top quartile of total centenarian rate (Montesanto et al., 2017). According to the official data available from the Italian National Institute of Statistics (ISTAT) at the time the COOL project was designed, in 2019 there were 386 individuals aged ≥99 (85% women) dwelling in the Genoa metropolitan area (ISTAT, 2021). To date, the Liguria region remains Italy’s longest-living region, with a median population age approaching 50, a centenarian rate of 59.4 per 100,000 residents as of January 1, 2025, and a semi-supercentenarians rate of 2.3 per 100,000 residents (ISTAT, 2025). In relative terms, Molise tops the list with approximately 61 centenarians per 100,000 residents. However, aside from this region, given its low absolute number of centenarians, Liguria, the most aged region in the country, holds a prominent position in terms of longevity. Among the provinces, Isernia has the highest concentration of centenarians (78.7), ahead of Nuoro (65.5), Siena and Gorizia (both 63.5). Three Ligurian provinces follow: Imperia (61.2), Genoa (61.1) and La Spezia (61.0). There were 35 semi-supercentenarians from Liguria over 105 years of age as of January 1, 2025, of which 20 in Genoa (ISTAT, 2025).

COOL is a not-for-profit, multicentre cohort study, involving a series of centenarians (aged >99) living in the Genoa area. The primary endpoint is the identification of genomic biomarkers associated with cognitive status of centenarians, using the cognitive profile as primary outcome measure. The output of the COOL cohort is a set of biological samples with associated medical and life history records which may have great value in a wide spectrum of ageing and well-being related research projects. The COOL study is currently in progress. Here we report the protocol of the study and the baseline characteristics of the new cohort of centenarians.

## Methods

### Study design

The COOL study is a not-for-profit, prospective, observational cohort study, aimed at assessing a series of centenarians living in the Genoa area (Liguria region, northwestern Italy).

The study was designed to investigate genomic biomarkers associated with cognitive healthy aging, using the cognitive profile as primary outcome measure. The relevant exposure is the genetic variability within the study cohort, as described by all type of genomic biomarkers that can be investigated as predictors. Sociodemographic variables, family history, clinical history, and their interactions with genomic variants, will populate different multivariate models to interpret the effect of genomic variations on cognitive measures. The study will not entail the investigation of disease-associated genomic variant, unless appropriate for clinical purposes.

The study protocol was designed on the basis of the current good clinical practice and the local health care practices determined by the national and regional health care regulations and procedures. The section on the clinical and neuropsychological assessment was partially aligned with the procedures adopted in the “Centenari a Trieste” study (Tettamanti and Marcon, 2018).

Family members and caregivers of the study participants were actively involved in the recruitment phase as well as during the conduction of the study. Participants and families will be involved in the dissemination phase, in collaboration with family doctors and practitioners of the nursing homes. The findings of the study will be reported in lay language, individually or in small groups during dedicated meetings. Beyond the scientific community, the dissemination plan will involve the diverse stakeholders involved in the COOL study (see **Figure 1**) – i.e. families, healthcare practitioners (physicians, nurses, psychologists), researchers, administrative officers and managers.

**Figure 1.**
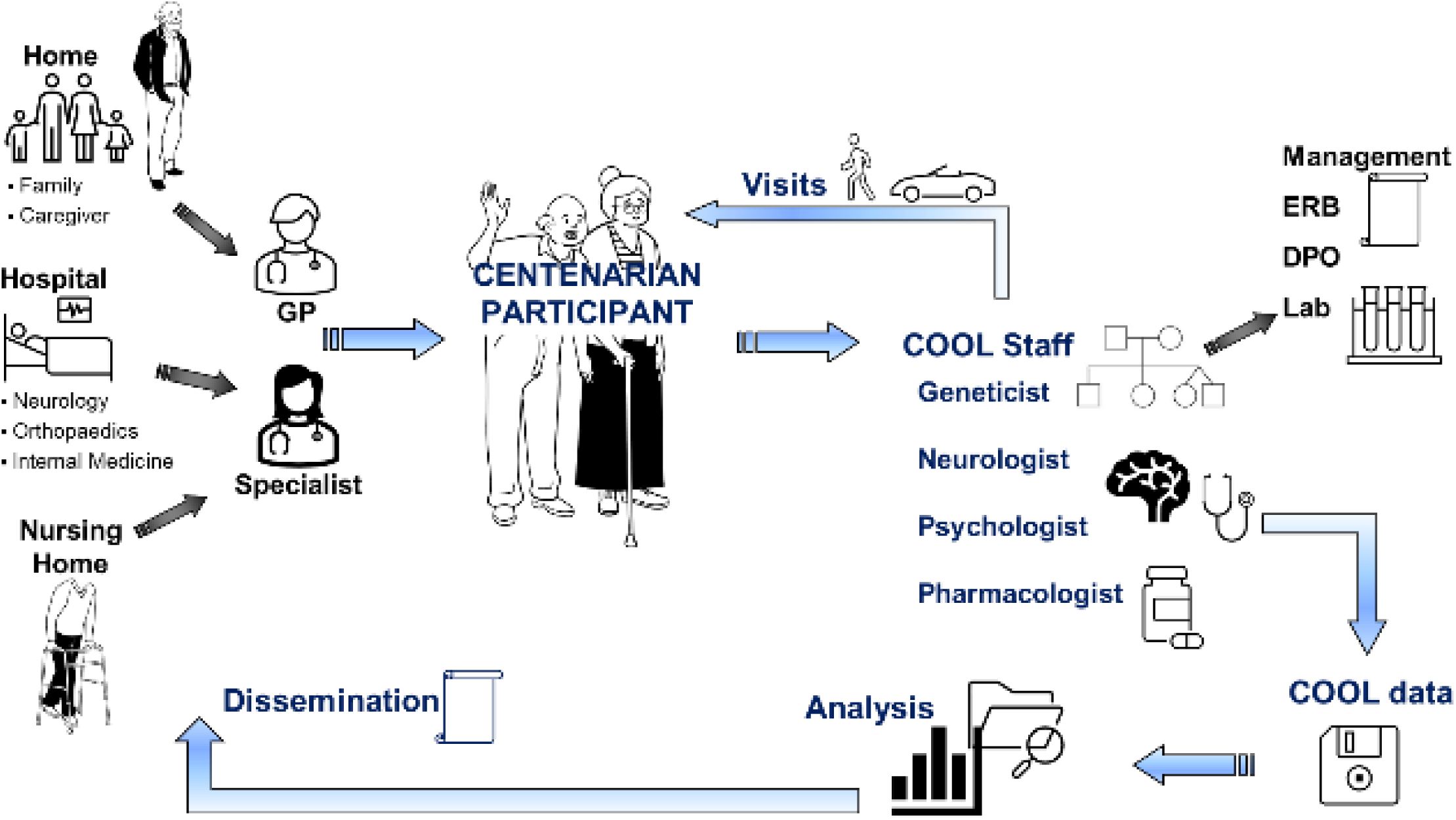
Workflow of the COOL study.

The study was approved by the Local Ethics Committee (Comitato Etico Territoriale, Regione Liguria, approval ID CER Liguria 91/2021).

### Participants

The primary enrolment area is the metropolitan area of Genoa, which roughly corresponds to the area under the responsibility of the local health care authority (“ASL3 Genovese”). At the time of the study design (2019) we estimated that approximately 300 individuals aged >100 were living in this area (data not shown). The target sample number (n=100 unrelated individuals) was set as a convenience sample size, based on the estimated population of centenarians in the catchment area and on a cautious estimation of the attrition rate. In order to extend the population of eligible participant, the cut-off age was set at ≥99 years of age. No formal power analysis was performed, according to the exploratory type of this study.

The eligible individuals were defined by the following inclusion criteria: i) age ≥99 years at the time of enrolment, regardless of the apparent cognitive status; ii) ability to sign the informed consent or, if the subject is not able to give consent, presence of the legal guardian or authorised delegate. Severe comorbidity, e.g., total blindness or clinically severe general conditions, and inability to provide the informed consent in absence of a legal guardian or authorised delegate, are the only exclusion criteria. Eligible individuals receive a full oral and written information, in presence of the caregiver and of the legal representative, if appropriate. Written informed consent is collected for all enrolled participants. The study was approved in its first version on March 2021, during the COVID-19 pandemic. The enrolment phase started in fact in late 2021, as the operative procedures in compliance with the ongoing pandemic could be put in place. In the first, pilot phase, the monocentric structure based on the Galliera Hospital was applied with the aim of evaluating the feasibility of the study.

The study protocol was amended in 2023 to implement to the following changes: i) a multicentric structure was adopted, to allow all hospitals serving the Genoa metropolitan area to enrol participants; ii) first-degree relatives (sibs and children) of centenarians are eligible for participation and can be assessed according to the same procedure. The latter amendment was grounded on the assumption that siblings of the first case are reliably close to the target age (≥99); both sibs and children can provide the study with important information on shared and not-shared genetic and environmental determinants. Furthermore, the enrolment period was extended until March 2028.

The minimum core of information includes sociodemographic data; past and recent medical history; past and recent drug consumption history; general examination (weight, height, BMI, heart rate, blood pressure, sarcopenia) and comorbidity through the Cumulative Illness Rating Scale (CIRS), a clinical tool used to quantify the elder individual’s burden of multimorbidity in a single index (Parmelee et al., 1995); we used the 1-5 scale, from no illness to extremely severe illness (Hudon et al., 2005). A summary of the information collected at the baseline visit is summarised in **Table 1**.

**Table 1.** Synopsis of the main assessments included in the COOL case report form.

| Domain assessed | Method and/or instrument | Content |
| --- | --- | --- |
| **Family History** | Interview, pedigree | Detailed family history, at least four generations (from grandparents to children) with ages and ages at death |
| **Ancestry** | Interview, pedigree | Geographical origin of parents and grandparents |
| **Marital status** | Interview, pedigree | Married / unmarried / divorced / widow (years) |
| **Children** | Interview, pedigree | Number, sex, current age |
| **Fertility** | Interview | Age at menarche and at menopause; number of pregnancies |
| **Spiritual status** | Interview | Religion, observant/not observant |
| **Education level** | Interview | Highest education level reached; years of education |
| **Occupational status** | Interview | Principal occupations in the past, current activities |
| **Retirement** | Interview | Age at retirement |
| **Leisure activities** | Interview | Past and present leisure activities |
| **Skills retained** | Interview | Visual ability, hearing, walking, incontinence, dysphagia |
| **Lifestyle** | Interview | Smoking, alcohol and drugs abuse, diet, sleep behaviour, motor activity |
| **Medical History** | Interview | Previous diseases, reasons and number of hospitalisations, reasons and number of surgeries, number of falls, current diseases |
| **Comorbidity status** | Cumulative Illness Rating Scale – CIRS (Parmelee et al., 1995) | Presence and severity of co-occurring medical conditions weighted for 14 main systems. Severity index: medium scores of domains 1-13. Comorbidity index: count of organ systems with score $\geq 3$. |
| **Vaccinations** | Interview | Covid-19, Flu, Herpes, Tetanus, Smallpox, Pneumococcus |
| **Pharmacotherapy** | Interview | Previous pharmacotherapy, current pharmacotherapy, autonomy in drugs administration, use of complementary medicine |

### Outcome Evaluations

The primary outcome measure is the cognitive profile of centenarians, as assessed by the means of a battery of validated neuropsychological tests. The core cognitive assessment at enrolment includes: i) the Mini-Mental State Examination (Folstein et al., 1975) (MMSE), which is the most used battery for elderly cognitive evaluation (Gualco et al., 2025) (validated in the Italian version by Measso et al., 1993); ii) the Clinical Dementia Rating scale (Hughes et al., 1982; Morris, 1997) (CDR), a widely used tool to stage cognitive abilities from no or mild impairment (score =0 or 0.5) to advanced dementia (score ≥4), by using a structured interview with both the patient and a reliable informant (family member or caregiver); iii) the Informant Questionnaire on Cognitive Decline (Jorm and Jacomb, 1989) (IQCODE), a short questionnaire administered to a proxy to assess cognitive decline in elderly people. In line with the 100-plus study (Holstege et al., 2018), we also record the researcher subjective impression of cognitive health. Moreover, the full examination battery include a subset of tasks taken from the CERAD battery (Welsh et al., 1994), that were used in a recent study on Italian centenarians (Tettamanti and Marcon, 2018). The battery of neuropsychological tests that were planned to assess the cognitive profile in the COOL study cohort is detailed in **Table 2**.

**Table 2.** Synopsis of the battery of neuropsychological tests administered to assess the cognitive profile of the COOL study participants.

| Domain | Assessment | Administered to | Ref. |
| --- | --- | --- | --- |
| *Cognitive functioning* |  |  |  |
| Global cognitive functioning | Researcher subjective impression of cognitive health |  | (Holstege et al., 2018) |
|  | Mini Mental State Examination (MMSE) | Participant | (Folstein et al., 1975; Measso et al., 1993) |
|  | Clinical Dementia Rating scale (CDR) | Participant, Caregiver | (Hughes et al., 1982; Morris, 1997) |
|  | Informant questionnaire on cognitive decline (IQCODE) | Caregiver | (Jorm and Jacomb, 1989) |
| Language | CERAD battery - Boston Naming Test (BNT) | Participant | (Morris et al., 1989; Tettamanti and Marcon, 2018) |
|  | CERAD battery - Semantic Fluency (Animal) | Participant | (Morris et al., 1989; Tettamanti and Marcon, 2018) |
| Memory | CERAD battery - 10 World List - Immediate | Participant | (Morris et al., 1989; Tettamanti and Marcon, 2018) |
|  | CERAD battery - 10 World List - Recognition | Participant | (Morris et al., 1989; Tettamanti and Marcon, 2018) |
|  | CERAD battery - 10 World List - Delayed Recall | Participant | (Morris et al., 1989; Tettamanti and Marcon, 2018) |
| Praxis | CERAD battery - Praxis | Participant | (Morris et al., 1989; Tettamanti and Marcon, 2018) |
|  | CERAD battery - Praxis Delayed Recall | Participant | (Morris et al., 1989; Tettamanti and Marcon, 2018) |
| Attention and Executive Functions | Trial Making Test A and B | Participant | (Greenlief et al., 1985) |
| Social Cognition | Reading the Mind in the Eyes Test - Short Form (RMET -SF) | Participant | (Chander et al., 2020) |
| *Psychiatric disorder* |  |  |  |
| Anxiety | Geriatric Anxiety Scale (GAS) - Short form | Participant | (Byrne and Pachana, 2011) |
| Depression | Geriatric Depression Scale (GDS) - Short form | Participant | (Herrmann et al., 1996) |
| *Functional Assessment* |  |  |  |
|  | Basic Activities of Daily Living (BADL – Barthel Index) | Caregiver | (Mahoney and Barthel, 1965) |
|  | Instrumental Activities of Daily Living (IADL) | Caregiver | (Lawton and Brody, 1969) |
|  | Advanced Assessment of Daily Living (AADL) | Caregiver | (Verghese et al., 2003) |
| Cognitive Reserve | Cognitive Reserve Index - questionnaire (CRI -q) | Participant / Caregiver | (Nucci et al., 2012) |

The secondary outcome measure is the cognitive decline of centenarians, assessed as the change in cognitive profile from the baseline to the follow-up visit. To assess cognitive decline with respect to the baseline evaluation, the neuropsychological examination is repeated after nine months from the first evaluation. This time interval was chosen to balance the high mortality rate of this cohort and the need to have sufficient time between the two evaluations in order to detect changes. The follow-up neuropsychological examination includes all the tasks performed at baseline.

### Procedures and workflow

COOL is an observational study. The participants do not undergo any experimental procedure. From the participant’s perspective, the comprehensive ascertainment outlined below does not substantially deviate from the procedures used in current clinical practice.

The **Figure 1** depicts the workflow of the whole project. Eligible participants are identified by healthcare practitioners, i.e. family doctors or home care specialists, or the relevant specialist if the centenarian was admitted to a hospital ward. The medical practitioner in charge is responsible for introducing the study to the participant and his/her family. The informed consent form is administered by a qualified investigator. An information letter is sent to the general practitioner. When participants are enrolled in a health and social residence the information is sent to the chief medical officer.

Centenarians were considered frail people; in this view they were not expected to move from their usual household for the purpose of the study. All the procedures are performed at the participants’ home or residence, or in the hospital where they were admitted. The history is collected in the presence of relatives or caregivers who help to provide information both verbally and through paper documentation. Enrolment and ascertainment were accomplished according to the current rules for the prevention of COVID-19.

Participants are assessed via a standard clinical assessment of the general health profile and undergo the battery of neuropsychological tests outlined above to ascertain the cognitive profile. The assessment includes collection of physiologic, familiar and pathologic history; general and neurological examinations; neuropsychological tests administration. The study does not include invasive procedures. The only minimally invasive procedure is blood sampling, which is performed, if accepted by the participant, by a trained physician or nurse. The **Table 2**. Synopsis of the battery of neuropsychological tests administered to assess the cognitive profile of the COOL study participants.**Table 2** reports a summary of the main assessment procedures.

Biological samples are processed and stored according to the guidelines for Good Laboratory Practice. Genomic and epigenetic biomarkers are being assessed retrospectively on all available biological samples. We do not report here the specific hypotheses that will be investigated by the means of deep genomic analysis. Relevant methods and findings will be described elsewhere.

### Data management

Personal information is treated in compliance with the UE Regulation 2016/679 (GDPR) and relevant Italian and Regional acts. Whenever possible, data on pathological conditions will be validated using information from existing health records, to minimise ascertainment errors. Data collected at participating centres are transferred in compliance with the current rules and best practice, preferably using hard copies. Data collected for the purpose of study are stored solely at the promoting centre. A unique pseudonym is assigned to each participant at enrolment and used in all subsequent procedures; the pseudonymisation key is securely stored at the promoter site. The Data Protection Impact Assessment document was developed at the study submission to the Ethics Committee and maintained in adherence to the current version of the study protocol. It did not reveal any critical area and was approved by the Data Protection Officer of the promoting centre.

### Biological samples

Venous blood (one or two EDTA tubes) was taken during a routine clinical evaluation. Sample aliquots were processed for DNA extraction using the MagDEA® Dx SV kit on the magLEAD 12gC system (Precision System Science Co, Ltd, Chiba, JP). Whole peripheral blood and DNA samples were stored at –20°C.

### Data analysis

For the present interim analysis, we considered records with a minimum set of data at enrolment, including: age (ascertained as date at enrolment minus date of birth); setting of assessment (household, nursing home, hospital); sociodemographic (place of birth, place of residence, education, main working activities); family history (ancestry of parents, number of children); recent clinical history and current medications; at least one measure of cognitive impairment (MMSE, CDR, researcher subjective impression). With regard to the primary outcome measure, participants were classified as suffering from definite severe dementia if scored either CDR ≥4 or MMSE ≤20 (Folstein et al., 1975). MMSE scores were compared across CDR categories using the Kruskal-Wallis test with correction for ties.

Quality control was carried out retrospectively on the dataset established in July 2025. Outcome variables were described by absolute and relative frequencies. Frequencies were estimated based on the number of assessed individuals for each variable. Descriptive statistics and diagrams were generated in Stata 16 (StataCorp LLC, USA). We report here the descriptive analysis of the baseline data at the time of enrolment (first visit).

## Results

### Participants

As of July 2025, we enrolled 88 participants with and without cognitive impairment, of whom 71 (81%) were women. The complete assessment (including interview, clinical examination, neuropsychological tests administration and biological samples harvesting) needed approximately two hours. Fractionated sessions were planned as needed.

The core set of demographic information was not recorded for 4 participants; therefore 84 individuals underwent the global assessment; 67 donated a biological sample (venous blood). Thirty-two subjects were followed up; 2 additional subjects who underwent the follow-up cognitive assessment, though not able to complete the examination at the first visit, were not included in the analysis of data at first visit. **Figure 2** reports the flow diagram of the enrolment and assessment process, as well as the reason for not completing the study protocol.

**Figure 2.**
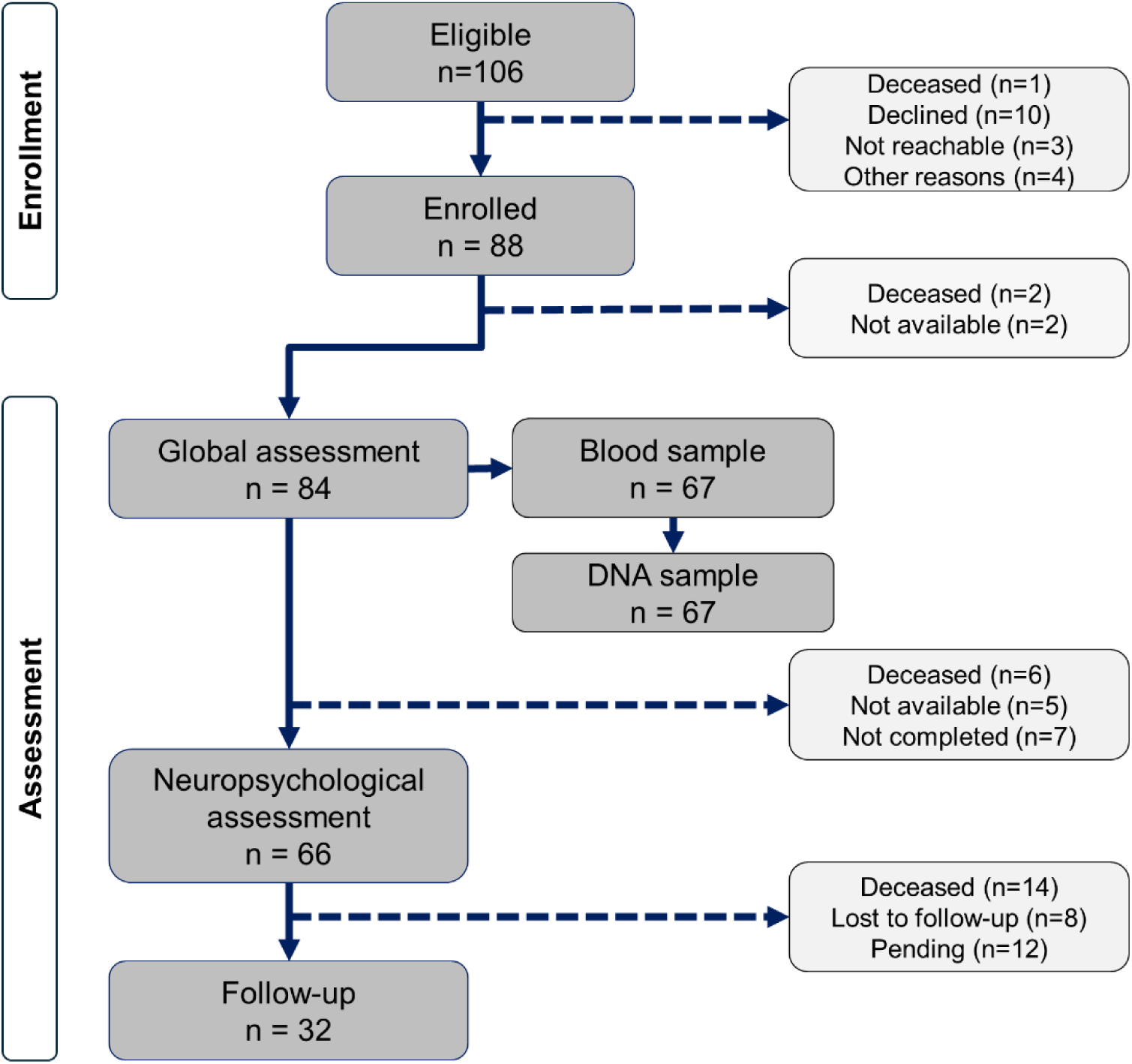
Flow diagram of the enrolment and assessment procedures, as of July 2025. The main reasons for not completing the relevant assessment phase are also reported.

In the cohort of 84 participants whose records passed the quality control, age ranged from 99 to 108 years (median: 100.56). At the last examination (i.e. including age at follow-up for 32 participants), the age range was 99-109 (median: 100.95). The full age distribution at enrolment and at the follow-up visit is reported in **Table 3**. Year of birth is comprised between 1913 and 1925, the most frequent being 1922 (25%).

**Table 3.** Age distribution of the COOL cohort (n=88). Age at last examination was defined as age at follow-up if completed (n=32), or age at enrolment for the remaining (n=56).

|  | n at | n at last |
| --- | --- | --- |
| age | enrolment | examination |
| 99 | 28 | 17 |
| 100 | 27 | 26 |
| 101 | 15 | 19 |
| 102 | 8 | 14 |
| 103 | 4 | 4 |
| 104 | 2 | 4 |
| 105 | 1 | 1 |
| 106 | 0 | 0 |
| 107 | 0 | 0 |
| 108 | 2 | 0 |
| 109 | 0 | 2 |

All participants were of Italian ancestry, as defined by having both parents of Italian origin. They were unrelated, with the exception of two siblings. All participants lived in the Genoa metropolitan area for most of their lives; 75% worked at least 10 years. At enrolment, 55% were living in a nursing home, 31% in their household with a relative or a caregiver, 13% alone. Participants attended a median number of 8 years of education. The mean severity index as measured by CIRS was 3 (SD 2.37).

### Cognitive status

Sixty-six participants were able to respond to the full interview and provided at least one measure of cognitive status at enrolment (CDR: n=64; MMSE: n=52); 2 additional individuals who did not undergo the full visit at enrolment were assessed only at the follow-up visit and were not included in the present analysis. The baseline assessment showed a wide distribution of cognitive status. CDR ranged from not impaired (CDR=0 assigned in 19 out of 64 participants, 29.7%) to severely demented, with 12 individuals classified as CDR ≥4 (18.4%). Those who completed the MMSE (n=52) showed a range of 3-30 (median 24, mean 22.79, SD 5.65). **Figure 3** depicts the MMSE score distribution as a function of the CDR categories in the participants who underwent both assessments (n=52). MMSE ranged from 23 to 30 (mean 26.73, SD 2.22), with significant decreasing trend over increasing CDR (Kruskal-Wallis test, χ²=41.465, 5 d.f., p<0.001). It appears that 30 participants were classified as either not impaired (CDR=0) or minimally impaired (CDR=0.5).

**Figure 3.**
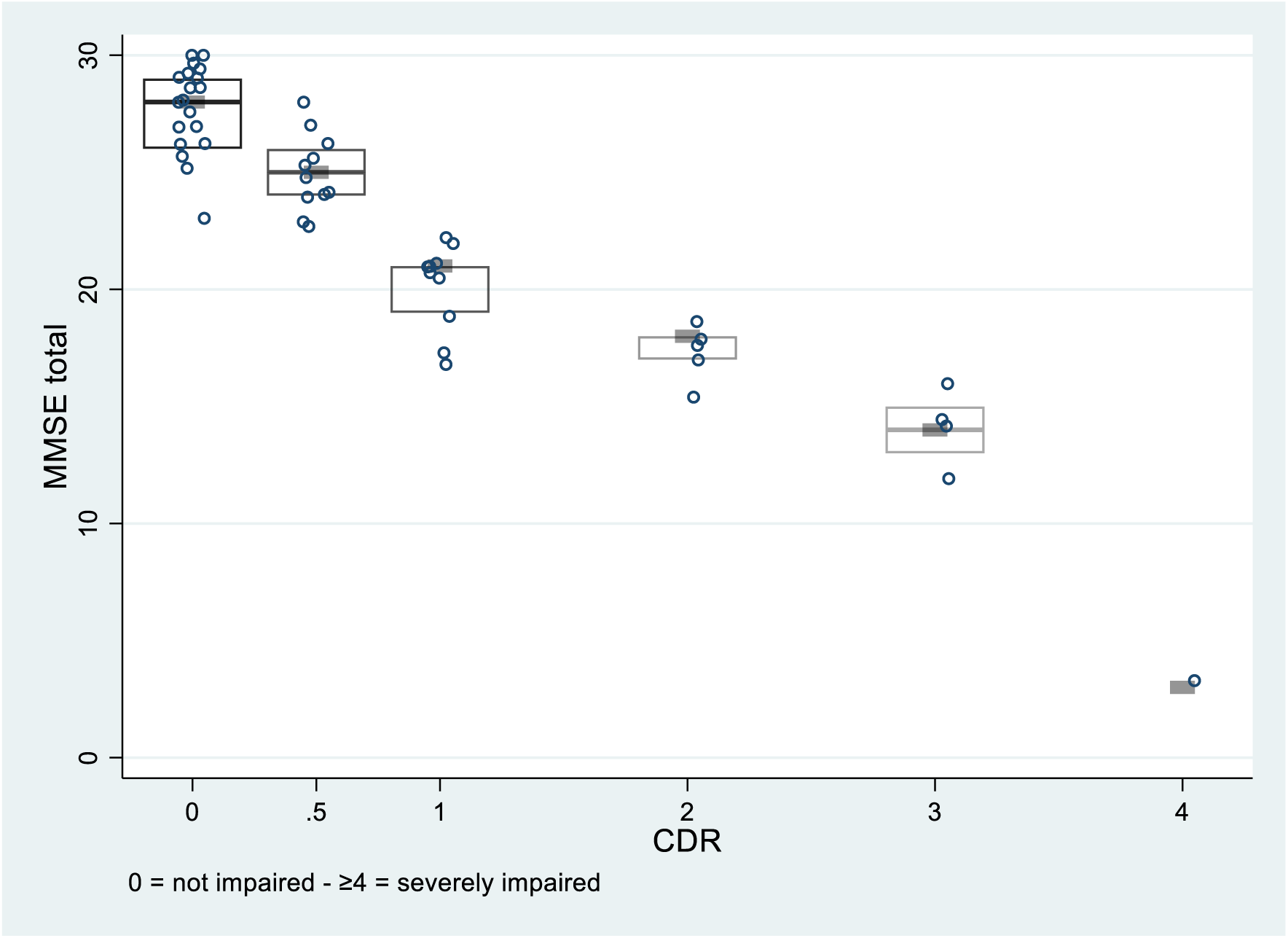
MMSE score distribution as a function of the CDR categories in individuals from the COOL cohort who underwent both assessments (n=52). The diagram shows the individual values, the interquartile range (box) and the median value (thick grey line).

Functional level, as operationalised by the Barthel index, ranged from 0 (totally dependent) to 100 (totally independent), median value 52.5 (mean 49.33, SD 33.08). **Figure 4** reports the frequency distribution of values of cognitive functioning (MMSE, CDR) and functional capacities (Barthel index), in individuals who were ascertained at enrolment. Further descriptive and inferential statistics will be reported elsewhere.

**Figure 4.**
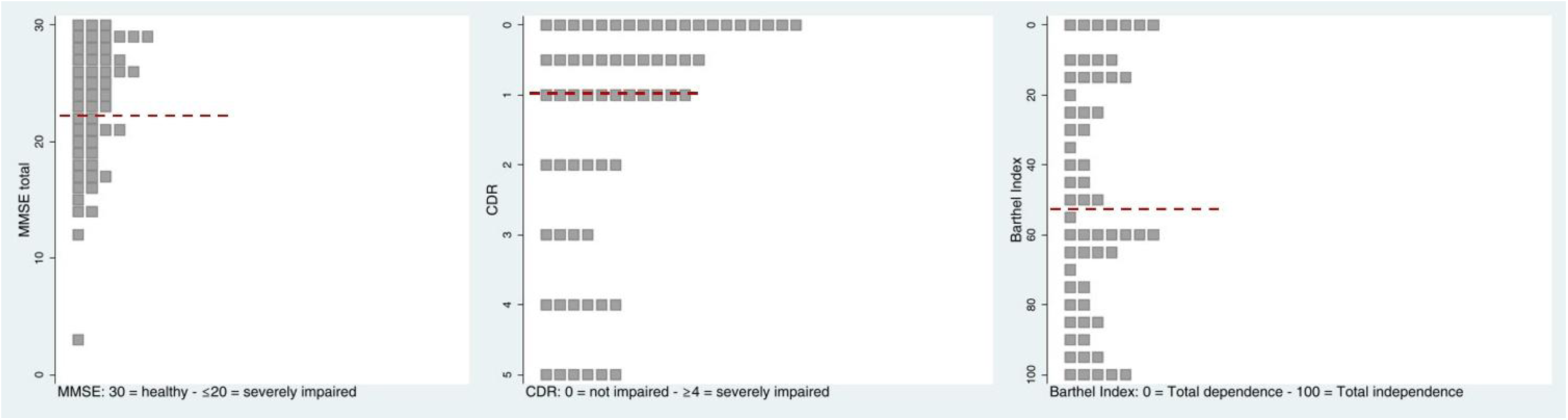
Frequency distribution of values observed in individuals from the COOL cohort for MMSE (n=52), CDR (n=64) and Barthel Index (n=60). The red dashed lines indicate median values. For clarity of comparisons, CDR and Barthel index scales were reversed to show better performance on top.

### Biological samples

Blood sampling could be carried out at any time after consent signing. Most participants consented to donate a residual blood sample for the purpose of the COOL study. To date, 67 blood samples were collected; DNA was extracted and stored.

## Discussion

It is still under debate whether centenarians and ultra-centenarians primarily avoided detrimental factors (“escapers”) or, to some extent, they were resilient to the same factors (“survivors”). This dichotomy applies also to cognitive decline during ageing, though mechanisms, i.e. resistance and resilience, supposedly contribute to healthy ageing (Andersen, 2020). Regardless of the underlying biological framework, selected cohorts of extremely aged individuals, namely aged 100 years or more, are invaluable models for studying favourable combination of constitutional and environmental protective factors (Borras et al., 2020).

The COOL study was designed as observational cohort study, aimed at elucidating the genetic determinants of cognitive health in the oldest population. More than one hundred individuals aged >99 years, and their families, were reached during the first approximately 3-year phase of recruitment. We enrolled 88 participants, close to the convenience target sample that was defined at the inception of the study. Most of them consented to provide detailed information on their personal and medical history. Detailed family history was collected from all participants. We gathered information on age at death and cognitive status, as ascertained from family history, regarding parents and grandparents of enrolled participants; their children and grandchildren often participated in the study interviews (the analyses from family history of the centenarians included in the COOL cohort will be described elsewhere).

The COOL study was originally designed in conjunction with the Centenarians in Trieste study (Tettamanti and Marcon, 2018). Genova and Trieste, both located in northern Italy, share a similar sociodemographic structure, as both cities present a high percentage of aged inhabitants, with a remarkable fraction of extreme aged individuals. The urban environment is also similar, as well as climate. Therefore, we aimed at developing a study protocol which allows to compare the two cohorts and eventually merge the respective data set.

Some insightful lessons were learnt during the accomplishment of the COOL study. We were aware that recruiting and assessing extremely aged individuals, in the real-world setting, are challenging tasks. When the protocol was designed, we cautiously overestimated the attrition rate. In fact, in our experience the identification of eligible centenarians was not a limiting factor. The network of practitioners, serving in diverse healthcare units located in the largest urban area of Genoa, was able to reach out a relevant number of eligible individuals. We believe that the close alliance between the candidate participants their families, and their physicians – whether the general practitioner, the neurologist or the geriatrician – favourably contributed to a trustworthy information and, in turn, facilitated the enrolment process. We also believe that the multidisciplinary approach adopted by including diverse professionals in the study team (i.e. neurologists, psychologists, pharmacologists, geneticists, in close connection with the data management and project management) was a fundamental added value. We thus demonstrated that the COOL study, was feasible in the current organisation of the healthcare practice. Notably, the eligible centenarians showed favourable attitude towards the study the study protocol, though the procedures were deemed demanding.

Since the organisational aspect was overcome, the first challenge of the COOL study was to demonstrate that variability in the primary outcome could be observed, in order to investigate the underlying determinants, and that biological samples could be collected, with no detrimental impact on the ongoing healthcare programme of individual participants.

Considering CDR and MMSE findings, both demented and non demented individuals were included in the interim COOL cohort. Among participants eligible to cognitive assessment and not severely impaired, cognitive performance ranged widely from unimpaired cognitive status to moderate impairment. The COOL study thus succeeded in enrolling a meaningful cohort of centenarians with diverse degree of cognitive impairment, from healthy cognitive profile to established dementia. In this regard, the COOL study differs from other research programmes on centenarians conducted in European countries. The seminal 100-plus study described by Holstege and coworkers (Holstege et al., 2018) recruited a large sample of Dutch centenarians who self-reported to be cognitively healthy. As a result, the 100-plus participants had relatively high cognitive profiles at baseline and retained hearing and visual abilities.

From the COOL participants’ perspective, the neuropsychological examination was the most demanding task. In continuity with previous studies (Brodaty et al., 2016; Legdeur et al., 2017), our recent systematic update confirmed that MMSE is the most used assessment tool in nonagenarians and centenarians (Gualco et al., 2025). The Digit Span, the Trail Making Test and the Semantic Fluency Test were frequently investigated tasks (see Gualco et al. (Gualco et al., 2025) and citations therein), due to their low complexity and the limited need of sensory skills to complete the test (Beker et al., 2019; Melikyan et al., 2019). Beyond cognitive status, the applicability of neuropsychological tests in the oldest old population is limited by vision and hearing deficits, fatigue, and tremors (Holstege et al., 2018; Whittle et al., 2007). Also the time needed is a major reason for not completing the neuropsychological assessment, as the oldest individuals may need more time to complete the tasks as compared to the elderly seniors (Melikyan et al., 2019). However, our literature search did not reveal agreed recommendations for overcoming these limitations (Gualco et al., 2025). Based on the existing knowledge we would suggest that a set of neuropsychological assessment tools should be developed to capture the wide distribution of cognitive performance in oldest individuals without dementia, and to remain feasible for centenarians with sensory and motor impairment, accounting for diversity of gender, education, and cultural and social background.

### Limitations

As we set only the lower limit of age at enrolment (≥99), and recruitment occurred over three years, the birth cohorts span approximately a decade. A potential influence of the Flynn effect (that is the improvement in the performance on cognitive tests over the years, according to (Flynn, 1999) could be argued. However, it may be noted that almost all COOL participants were born after 1918, as life conditions in Europe increasingly improved and mortality decreased (Robine and Cubaynes, 2017); all were adolescents or young adults during the World War II.

According to the real-world setting and the simple inclusion criteria (i.e. age ≥99 and informed consent, regardless of the apparent cognitive status), participants were not systematically selected. Most subjects were recruited through their family doctor or the geriatrician in charge in the institutional household; in the latter case they were often not autonomous. Thus, some degree of ascertainment bias is likely.

In fact, the study was designed with the aim of assessing a heterogeneous sample of centenarians. Conversely, the restricted catchment area, that is the urban area of Genoa, was chosen to guarantee that homogeneous healthcare services were offered to the centenarians along most of their lives. Moreover, participants in the COOL study grew up, worked, and reached advanced age, while sharing the same social and environmental context. The families settled in Genoa during the twentieth century originated from diverse Italian regions, reflecting sustained internal migration toward the city during industrialization (Poli and Candura, 2021), though all belonging to the ancestral populations of the modern European area (Montesanto et al., 2017). The admixture of individuals with diverse Italian origins may be seen as a weakness. However, genetic diversity underlying the phenotype of interest is inherent to the study design, as COOL is aimed at capturing all type of genomic variations that can be detected in the study sample. Furthermore, the presence of discrepant genetic background will be checked by the means of genomic control, a method which discriminates population stratification or cryptic relatedness in genetic association studies, with no need of family analysis (Zheng et al., 2006). A fraction of the cohort could not be assessed for the full set of demographic, medical and cognitive measures. After enrolment, some individuals deceased; in other instances, the general condition worsened rapidly and hampered the examination. In compliance with the observational, real-world design of the study, we refrained from reiterating the attempts to reach the participants who missed the evaluation or follow-up. As a consequence, the final attrition rate prevented us to complete the cognitive assessment and blood sampling in the large majority of participants. Nonetheless, more than half of the eligible individuals were fully assessed. Given the intrinsic design of the study and the target population – i.e. centenarians – we believe that these limitations are not significantly affecting the actionability of the COOL cohort.

### Perspectives and future aims

The development of models to interpret the interplay between protective and detrimental factors in extreme ageing is not a mere scientific speculation, though of utmost interest. Given the burden of cognitive impairment on the individuals, their families and healthcare services, healthy cognitive ageing is a major public health priority. The identification of a subset of individuals predicted to be particularly vulnerable to cognitive decline may enable targeted interventions. Preventive measures in healthy individuals, and therapeutical intervention in the early stage of decline, should be the priority. As an example, the recently developed anti-amyloid drugs may be helpful to slow the disease progression in patients who are close the onset of symptoms; their use, though, is constrained by serious limitations and adverse effects (Thambisetty and Howard, 2023) and should be avoided in individuals who are not predicted to experience a severe disease progression (Fox et al., 2025). Other innovative approaches, such as those based on p75 neurotrophin receptor signalling pathways (Shanks et al., 2024), may lead to effective protective drugs in the near future.

As the expected lifespan at birth has been rapidly increasing, and the number of extreme aged individuals is raising correspondingly, the scientific and clinical communities should focus on the length of healthy life, and how to improve it in an equitable manner for all citizens. The updated Lancet Commission report (Livingston et al., 2024) underlined that prevention approaches should be put in place early and, remarkably, emphasised that the actions to decrease risk factors exposure should be carried forward throughout life (i.e., the longer, the better). Moreover, it was reported, for the first time, that risk can be modified even in people with increased genetic liability to dementia (Livingston et al., 2024). This novel framework for actions towards the reduction of ill health life particularly urges the universalistic health systems, namely in European countries and specifically in Italy, to design interventions that may substantially improve equity. As Liguria is the region with the oldest population in Italy, the comprehensive characterization of long-lived subjects is crucial to develop knowledge-based health policies. The multidimensional dataset on the cohort of centenarians will constitute a scientific standpoint to explore novel research questions and to establish fruitful partnerships with existing initiatives.

According to the ultimate goal of the COOL study, the body of data on the phenotype of the study cohort, namely the multimodal assessment of the cognitive profile, will allow us to design the eventual analyses of genetic determinants of the cognitive health status in the oldest old population. Genomic analyses (currently in the pilot phase) will employ the latest sequencing technologies and will be informed by the newest findings on the genetic architecture of extreme ageing (Tesi et al., 2026). The full data set collected from the general and neuropsychological assessment will populate the inferential analyses of the COOL cohort.

The COOL study was grounded in the open science paradigm. The investigators recognised the value of transdisciplinary collaboration and of knowledge sharing. Cognitive health in extremely old individuals represents a rare phenotype, and its investigation is fostered by broad collaborative networks. The comparison of cohort characteristics from diverse cohorts may provide insightful hints to further explore the determinants of healthy cognitive ageing. In this context, the COOL study dataset and biological samples will be made available to the scientific community on a collaborative basis.

## Conclusions

After the first phase of the COOL study implementation, a large phenotype data set was established from a cohort of centenarians who lived in the Genoa urban area. The relevant biological samples were collected, and DNA samples are available for molecular analyses. The multidisciplinary, local network of scholars and practitioners was an added value to accomplish the study procedures.

It is worth to underline that the COOL study was not designed to explore longevity *per se* – that is, how and why the centenarians in Genoa became centenarians. Conversely, the research question investigated in the COOL study addresses the hypotheses concerning the constitutional factors which determined, in those who reached 100 years of age, the retention of favourable cognitive health, as opposed to the centenarians whose cognitive status declined.

The interim findings demonstrated that participants exhibit a wide distribution of the cognitive profile, as formally assessed by the means of a panel of test. A remarkable proportion of participants is cognitively preserved. Therefore, we could meet the assumption that a series of centenarians from the real-world setting shows an actionable distribution of the cognitive profile. The molecular underpinnings of successful ageing could inform health policy strategies, ultimately contributing to increased healthy lifespan in the general population.

## Data Availability

The data that support the findings of this study are available from the corresponding author upon reasonable request.

## Acknowledgements

We thank the participants and their families for the generous participation into the study. The coauthors collectively listed as “COOL study investigators” contributed to the participants’ enrolment and assessment. We gratefully acknowledge for the valuable assistance the Research Office of the Galliera Hospital, especially Dr. Alessandra Argusti (currently at San Martino Hospital) and Dr. Alessandra Seri.

## Collaborators

The COOL study Investigators, who should be regarded to as co-authors, are: Maria Gabriella Poeta, Giancarlo Antonucci (EO Ospedale Galliera); Pietro Astuni, Fabio Piras (Department of Internal Medicine, Pharmacology & Toxicology Unit, University of Genoa); Raffaella Boi, Fortunato Lugarà, Nives Parodi (Azienda Tutela Salute Liguria); Adriana Agnese Amaro, Giammarco Baiardi, Paola Del Sette, Zeinab El Rashed, Enrico Pedemonte, Silvia Pesenti, Ulrich Pfeffer, Davide Sassos (IRCCS Azienda Ospedaliera Metropolitana Genovese); Diego Artuso, Dario Olobardi, Paola Spatera (Ospedale Evangelico Internazionale); Pierclaudio Brasesco, Arta Cakoni (Medicoop Liguria); Paola Fuliano, Rosella Giusto (Istituto Don Orione – Quarto Castagna); Tommaso Biondani (RSA Casa Serena); Rita Bartolozzi (RSA Coronata); Laura Norelli (RSA Opera Pia Causa):; Simona Badino, Mauro Pruzzo, Alba Cecilia Rodriguez Gavilanes (RSA Villa Duchessa di Galliera); Filippo Bussi (Residenza Sunflower)

## Authors’ contribution

EDM: conceptualization (lead); formal analysis (lead); funding acquisition; supervision (lead); writing – original draft (lead); writing – review and editing. CG: data curation; formal analysis; investigation; project administration. EMu: data curation; investigation. NR: data curation; investigation; project administration. CMS: funding acquisition; resources; supervision. LC: investigation; resources. UT: resources; supervision. CI and LM: investigation; project administration. FB: methodology; supervision. EMa: investigation. MS: data curation; formal analysis; investigation; writing – review and editing. FM: supervision; writing – review and editing. EZ: investigation. GF: supervision. MM and NS: resources. AS: methodology; supervision. GM and MT: conceptualization; writing – review and editing. COOL study Investigators: investigation. MDS: conceptualization; methodology; resources; supervision; writing – review and editing.

## Patient consent for publication

All participants provided informed consent for the purposes of the present study, including publication of findings.

## Provenance and peer review

not commissioned; externally peer reviewed.

